# Cholesterol distribution among adults in Nigeria: A descriptive cross-sectional study in a rural tertiary hospital

**DOI:** 10.1101/2024.04.23.24306215

**Authors:** Danny Asogun, Ralph K Akyea, Kingsley C Osuji, Nathaniel O Imafidon, Nadeem Qureshi, Barbara Iyen

## Abstract

**Background:** There is an increasing burden of cardiovascular disease in the Nigerian population, but very little known about the cholesterol distribution within subgroups of the general population.

**Methods:** A cross-sectional study of adults was done in a tertiary hospital in the Southern region of Nigeria. Using results of blood tests over an 8-month period, the cholesterol distribution and prevalence of dyslipidaemias were determined.

**Results:** A total of 46.8% of patients had raised total cholesterol (mean total cholesterol 198.93mg/dl (SD 57.81)), 14.6% had raised low-density lipoprotein cholesterol (LDL-C) (mean LDL-C 120.83mg/dl (SD 51.62)), and 27.9% of patients had raised triglycerides (mean triglycerides 128.83mg/dl (SD 75.26). Women had higher mean values than men, of total cholesterol (203.3mg/dl vs 193.4mg/dl), LDL-C (125.1mg/dl vs 115.5mg/dl), and non-HDL cholesterol (148.4mg/dl vs 141.1mg/dl). The 95^th^ percentile values of total cholesterol and LDL-C in the general population were 294mg/dl (7.6mmol/L) and 205.7mg/dl (5.3mmol/L) respectively.

**Conclusion:** Hypercholesterolaemia is highly prevalent in the Nigerian general population, with a higher prevalence in women compared to men. The 95^th^ percentile identified is likely to represent the thresholds associated with elevated risk of atherosclerotic cardiovascular disease risk within this population.

**What is already known on this topic:** There is a rising prevalence of cardiovascular disease (CVD) and CVD-related deaths in Nigeria, and a pressing need for targeted strategies and interventions to reduce this burden. However, there is lack of robust evidence on cholesterol distribution within subgroups of the Nigerian population.

**What this study adds:** This study described the distribution of cholesterol concentration in Nigerian adults by sex and age groups, and also determined the cholesterol 95^th^ percentile thresholds which are likely to be associated with elevated risks of atherosclerotic cardiovascular disease risk within this population

**How this study might affect research, practice or policy:** Evidence from this study will aid appropriate targeting of interventions to reduce the burden of hypercholesterolaemia and consequent risk of atherosclerotic cardiovascular disease.

## Introduction

Cardiovascular diseases (CVD) are a major cause of morbidity and the leading cause of mortality globally (1). Ethnic and regional differences exist in the prevalence of cardiovascular disease and CVD-related death (2), with over three-quarters of global CVD-related deaths occur in low- and middle-income countries (3). Hypercholesterolaemia, particularly raised low-density lipoprotein (LDL) cholesterol, is a causal risk factor for atherosclerotic cardiovascular disease (4) and remains the primary treatment target for clinical guidelines for both primary and secondary prevention (5, 6).

Despite the high prevalence of hypercholesterolaemia (7, 8), as well as the increasing burden of cardiovascular disease in the Nigerian population (9), there is very little known about the blood cholesterol distribution within the Nigerian population. Determining and understanding the distribution of cholesterol and other lipids such as LDL-cholesterol, triglycerides, and non-high-density lipoprotein (HDL) cholesterol in the Nigerian population will support targeted strategies and interventions to reduce the burden of cardiovascular disease and CVD-related mortality.

This study aimed to describe the mean values of total cholesterol, LDL-cholesterol, and non-HDL cholesterol among different sex and age groups of adults in the Southern part of Nigeria.

## Methods

A descriptive cross-sectional study was conducted in Irrua Specialist Teaching Hospital (ISTH), a teaching hospital in Edo State, Southern Nigeria, using retrospective records of all adult patients who had blood tests for cholesterol level estimation at the chemical pathology laboratory of the hospital from April 2022 to November 2022. We described the distribution of blood cholesterol (LDL-cholesterol, non-HDL cholesterol, and triglyceride levels) in the general population of patients who were treated in this hospital. Using the ISTH laboratory range of normal values, (total cholesterol <200mg/dl (or <5.17mmol/L), LDL-cholesterol upper limit of 159mg/dl (or 4.11mmol/L), and triglycerides <150mg/dl (or <1.69mmol/L), we also determined the prevalence of dyslipidaemia within subgroups of the population. All analyses were conducted using Stata SE version 17 statistical package. Ethical approval for this study was granted by the ISTH Human Research and Ethics Committee (approval ID ISTH/HREC/20222609379). Medical records of patients were accessed retrospectively on 26 February 2023 and the study authors only had access to the fully anonymised records.

## Results

There were 1,309 patients in the study which comprised 577 men (44.1%) and 732 women (55.9%). The mean age of patients was 46.7 (SD 16.2) years (48.4 years for men; 45.4 years for women).

Table 1 shows the mean values of total cholesterol, LDL-cholesterol, triglycerides, and non-HDL cholesterol overall, as well as separate values for men and women stratified by 10-year age groups. Women had higher overall mean values of total cholesterol (203.3mg/dl vs 193.4mg/dl), LDL-C (125.1mg/dl vs 115.5mg/dl), and non-HDL cholesterol (148.4mg/dl vs 141.1mg/dl) compared to men. These values were higher in women than men in all 10-year age groups except the 30–39-year age group where men had higher mean values, than women, of total cholesterol (200.3mg/dl vs 196.0), LDL-C (125mg/dl vs 119.1mg/dl), and non-HDL cholesterol (148.8mg/dl vs 142.7mg/dl). Although, overall, mean triglyceride levels were higher in men than women, triglyceride levels were higher in women than men in most 10-year age groups except the 40–49-(153.9mg/dl vs 119.4mg/dl) and the 60–69-year age groups (133.6mg/dl vs 121.9mg/dl) where men had higher levels than women.

**Table 1.** Baseline and lipid characteristics of the study population.

|  | Total (n=1,309) | Men (n=577) | Women (n=732) |
| --- | --- | --- | --- |
| **Mean age, mean(SD)** | 46.7 (16.2) | 48.4 (17.3) | 45.4 (15.2) |
| **Total cholesterol in mg/dl, mean (SD)** | 198.93 (57.81) | 193.36 (61.11) | 203.33 (54.71) |
| TC 50th centile in mg/dl (IQR) | 198.93 (195.80-202.06) | 193.36 (188.37-198.35) | 203.33 (199.36-207.29) |
| TC 90th centile in mg/dl (IQR) | 273.02 (268.79-277.24) | 271.67 (264.94-278.40) | 273.44 (268.09-278.79) |
| TC 95th centile in mg/dl (IQR) | 294.02 (289.21-298.82) | 293.87 (286.22-301.52) | 293.32 (287.24-299.40) |
| TC 99th centile in mg/dl (IQR) | 333.41 (327.38-339.44) | 335.51 (325.91-345.12) | 330.60 (322.97-338.24) |
| **Total cholesterol by age group** |  |  |  |
| 18-29 years (n=265) | 196.30 (57.80) | 192.98 (66.19) | 198.80 (50.64) |
| 30-39 years (n=153) | 197.56 (71.70) | 200.32 (94.83) | 195.96 (54.58) |
| 40-49 years (n=303) | 201.73 (54.26) | 193.82 (55.05) | 207.36 (53.12) |
| 50-59 years (n=271) | 198.37 (52.84) | 191.21 (50.63) | 202.90 (53.85) |
| 60-69 years (n=198) | 202.58 (53.83) | 197.11 (54.61) | 209.13 (52.44) |
| 70-79 years (n=93) | 201.91 (65.25) | 192.68 (52.36) | 212.65 (76.87) |
| >=80 years (n=26) | 168.81 (53.10) | 162.83 (11.52) | 182.25 (62.98) |
| **LDL cholesterol in mg/dl, mean (SD)** | 120.83 (51.62) | 115.47 (48.00) | 125.05 (53.97) |
| LDL-C 50th centile in mg/dl (IQR) | 120.83 (118.03-123.63) | 115.47 (111.56-119.39) | 125.05 (121.14-128.96) |
| LDL-C 90th centile in mg/dl (IQR) | 186.99 (183.21-190.76) | 176.98 (171.69-182.27) | 194.22 (188.94-199.50) |
| LDL-C 95th centile in mg/dl (IQR) | 205.74 (201.45-210.03) | 194.42 (188.41-200.43) | 213.83 (207.83-219.83) |
| LDL-C 99th centile in mg/dl (IQR) | 240.92 (235.54-246.31) | 227.13 (219.58-234.67) | 250.61 (243.08-258.14) |
| **LDL-cholesterol by age group** |  |  |  |
| 18-29 years (n=265) | 118.68 (42.26) | 118.36 (49.17) | 118.93 (36.35) |
| 30-39 years (n=153) | 121.24 (58.99) | 124.95 (81.82) | 119.09 (40.78) |
| 40-49 years (n=303) | 124.29 (71.42) | 112.44 (44.81) | 132.72 (84.58) |
| 50-59 years (n=271) | 119.02 (39.66) | 112.11 (41.36) | 123.39 (38.03) |
| 60-69 years (n=198) | 122.31 (37.37) | 117.01 (38.03) | 128.67 (35.76) |
| 70-79 years (n=93) | 123.59 (45.80) | 117.10 (37.32) | 131.14 (53.48) |
| >=80 years (n=26) | 97.73 (35.45) | 94.72 (31.70) | 104.5 (44.40) |
| **Triglycerides in mg/dl, mean (SD)** | 128.83 (75.26) | 133.32 (84.67) | 125.30 (66.75) |
| **Triglycerides by age group** |  |  |  |
| 18-29 years (n=265) | 124.06 (72.87) | 121.87 (74.76) | 125.71 (71.62) |
| 30-39 years (n=153) | 129.18 (79.95) | 127.27 (80.53) | 130.28 (80.02) |
| 40-49 years (n=303) | 133.74 (87.02) | 153.91 (116.03) | 119.38 (54.16) |
| 50-59 years (n=271) | 127.99 (64.49) | 127.58 (63.05) | 128.25 (65.57) |
| 60-69 years (n=198) | 128.25 (75.14) | 133.55 (79.32) | 121.89 (69.70) |
| 70-79 years (n=93) | 132.17 (63.11) | 131.42 (70.74) | 133.05 (53.72) |
| >=80 years (n=26) | 119.58 (71.75) | 118 (53.25) | 123.13 (107.15) |
| **Non-HDL cholesterol in mg/dl, mean (SD)** | 145.16 (48.64) | 141.07 (2.27) | 148.39 (1.60) |
| **Non-HDL by age group** |  |  |  |
| 18-29 years (n=265) | 142.40 (47.08) | 139.18 (56.95) | 144.83 (37.99) |
| 30-39 years (n=153) | 144.96 (62.33) | 148.82 (84.44) | 142.73 (45.31) |
| 40-49 years (n=303) | 147.17 (44.98) | 143.19 (46.65) | 149.99 (43.66) |
| 50-59 years (n=271) | 144.54 (48.52) | 137.40 (57.11) | 149.05 (41.76) |
| 60-69 years (n=198) | 147.95 (43.94) | 143.26 (45.22) | 153.58 (41.89) |
| 70-79 years (n=93) | 149.20 (50.55) | 142.36 (41.87) | 157.16 (58.58) |
| >=80 years (n=26) | 121.96 (38.15) | 118.78 (35.42) | 129.13 (45.46) |

Using the ISTH laboratory range of normal values, a total of 46.8% of patients (41.8% of men and 50.7% of women) had raised total cholesterol, 14.6% (12.8% of men and 16% of women) had raised low-density lipoprotein (LDL) cholesterol, and 27.9% of patients (28.8% of men and 27.2% of women) had raised triglyceride levels. The 95^th^ percentile values of total cholesterol and LDL-cholesterol in this study population were 294 mg/dl (or 7.6mmol/L) and 205.7 mg/dl (or 5.3mmol/L) respectively.

**Figure 1.**
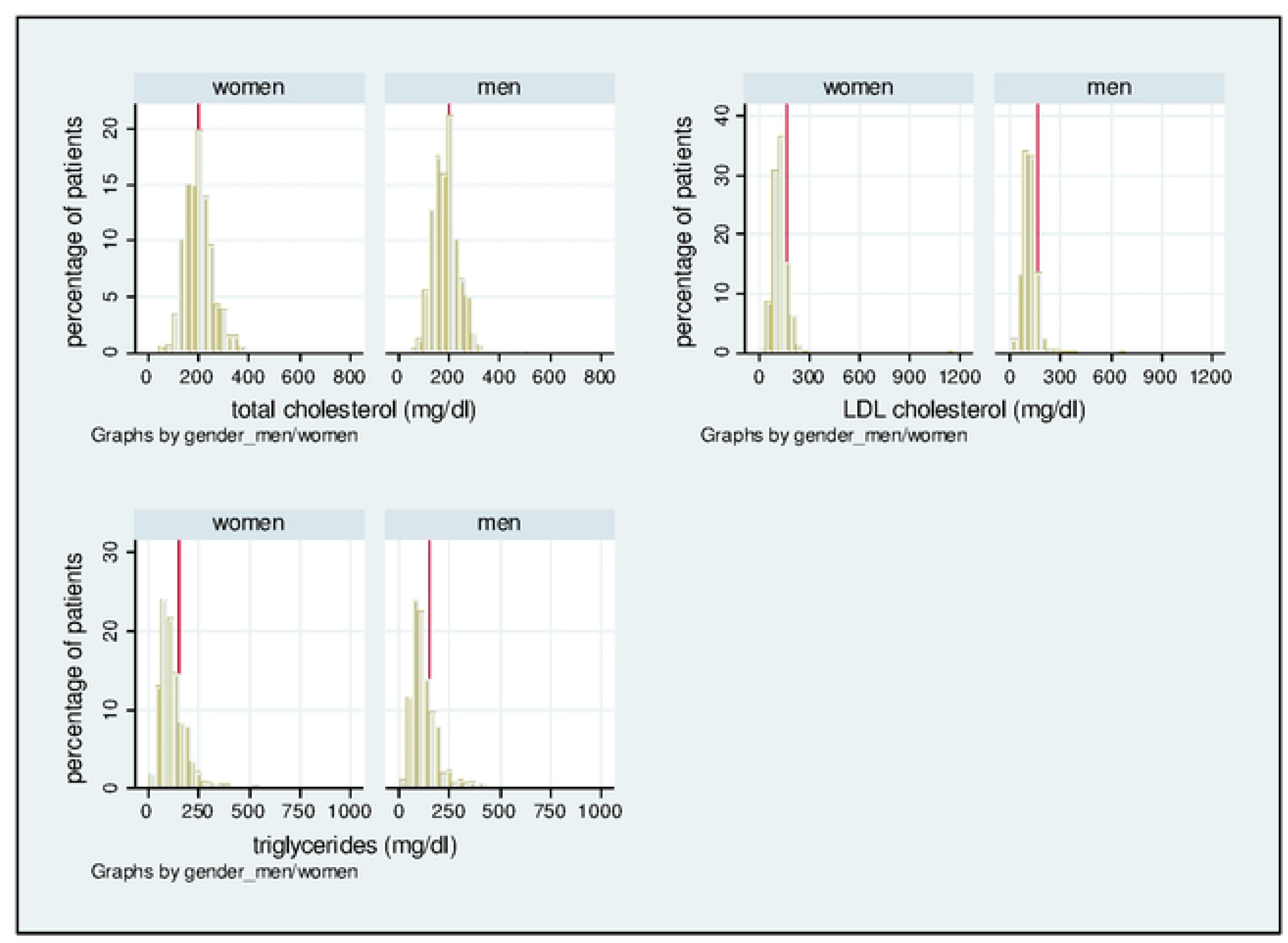
Distribution of total cholesterol, LDL-cholesterol, and triglycerides in men and women. *Vertical line in plots show the normal cut-off values for the different lipid measures *The heights of the bars are scaled so that the sum of their heights equals 100

## Discussion

In this descriptive cross-sectional study of adults in the Southern region of Nigeria, mean values of total cholesterol, LDL-C and non-HDL cholesterol were higher in women than men. This was consistent across all 10-year age groups except the 30–39-year age group where the values were higher in men than women. Triglyceride levels were higher in women than men in most 10-year age groups except the 40–49-, and the 60–69-year age groups where values were higher in men. There was raised total cholesterol (200mg/dl or higher) in 46.8% of the study population, 14.6% had raised LDL-cholesterol (above 159 mg/dl), and 27.9% had raised triglyceride levels (150mg/dl or higher). The 95^th^ percentile threshold values of total cholesterol and LDL-cholesterol within this population were 294mg/dl and 205.7mg/dl respectively.

### Comparison with existing literature

Our study finding of a higher prevalence of hypercholesterolaemia in women compared to men is similar to findings reported in a systematic review which estimated a 42% prevalence of hypercholesterolaemia in women (95%CI 23%-63%) and a 38% prevalence in men (95%CI 20%-58%) in Nigeria (8).

### Strengths and limitations

To our knowledge, this is the first study to describe the distribution of cholesterol concentration in Nigerian adults by sex and age groups. It is also the first study to determine the 90^th^, 95^th^ and 99^th^ percentile threshold of these cholesterol measures, in men and women, which are likely to represent the thresholds associated with elevated risk of atherosclerotic cardiovascular disease risk within this population. We acknowledge a few limitations of the study. Our study presents cholesterol results of adults who undertook blood tests at the tertiary hospital over an 8-month period. These would include patients referred for blood tests as part of the clinical work-up for the management of cardiac diseases including hypertension, stroke, obesity, diabetes, renal diseases, other conditions which may be associated with raised cholesterol, routine medical examination, as well as individuals undergoing screening to obtain medical certificates of fitness for new employment or travel purposes. While those studied are likely representative of adults in the Southern region of Nigeria in terms of demographics, lifestyle and diet, we have not included data from certain groups of the population who are apparently healthy or the hard-to-reach, so there is a small risk of selection bias.

### Clinical Implication

Our study findings highlight the burden of hypercholesterolaemia within the general population of Nigerians and support evidence from previous studies which report a disproportionately greater burden of hypercholesterolaemia in women. This underlines the importance of clinical and public health strategies which are appropriately targeted, to reduce the burden of hypercholesterolaemia and consequent risk of atherosclerotic cardiovascular disease. The 95^th^ and 99^th^ percentile thresholds of total cholesterol and LDL-C identified in this study likely indicate the population threshold at or above which patients are at significant risk of atherosclerotic cardiovascular disease, as well as a high possibility of inherited hypercholesterolaemias such as familial hypercholesterolaemia. Importantly, there is the need for large scale studies to determine the appropriate total cholesterol or LDL-C threshold which is associated with elevated risk of atherosclerotic cardiovascular disease in the general population of Nigerians.

## Data Availability

Data cannot be shared publicly, but may be made available on request from the ISTH Human Research and Ethics Committee for researchers who meet the criteria for access to confidential data.

